# Assessing the reach of the IMAM program in a remote setting: Evidence from Bajura district, Nepal

**DOI:** 10.1101/2025.08.26.25334514

**Authors:** Lila Bikram Thapa, Sujay Nepali, Roshna Maharjan, Ashish Timalsina, Melkamnesh Alemu Nigussie, Phulgendra Prasad Singh, Manisha Katwal

## Abstract

**Background:** Wasting remains a major public health concern in Nepal, contributing to preventable morbidity and mortality among children under five. The Integrated Management of Acute Malnutrition (IMAM) program has been implemented in Bajura district since 2014/15, yet evidence on its coverage and accessibility has been lacking. This study assessed program coverage, identified barriers and enablers, and proposed strategies to improve reach and effectiveness.

**Methods:** A Semi-Quantitative Evaluation of Access and Coverage (SQUEAC) approach was employed in three stages. Stage I combined routine HMIS data review with qualitative data from key informant interviews, focus group discussions, and direct observations to identify barriers and boosters. Stage II tested the hypothesis that coverage was higher near Outpatient Therapeutic Care Centres (OTCCs) and lower in distant areas, using active and adaptive case-finding. Stage III conducted a wide-area survey, applying Bayesian analysis to estimate coverage.

**Results:** The assessment identified 14 boosters, including OTCC expansion and integration of SAM screening into growth monitoring and immunization services, and 25 barriers, notably RUTF stockouts, inadequate health worker capacity, and low community awareness. Coverage was estimated at 23.5% (95% CI: 14.5–36.0%), well below the SPHERE standard of ≥50% for rural settings. Point coverage was 11.53%, indicating very limited reach at any given time. Female children constituted the majority (69%) of uncovered cases, suggesting possible gender-related disparities.

**Conclusion:** IMAM coverage in Bajura is substantially below international benchmarks, constrained by both service delivery gaps and socio-cultural barriers. Strengthening health worker capacity, ensuring uninterrupted RUTF supply, enhancing community engagement, improving data systems, and addressing gender-related inequities are critical to expanding access and improving treatment outcomes.

## Introduction

Acute malnutrition remains a major global health challenge, contributing to nearly half of all deaths among children under five years, especially in low- and middle-income countries [1]. Children with severe wasting are approximately 11 times more likely to die from common childhood illnesses than healthy children [2]. In Nepal, despite some progress, the burden of undernutrition remains high. According to the 2022 Nepal Demographic and Health Survey, 25% of children under five are stunted, 8% are wasted, and 19% are underweight [3]. These figures reflect a persistent gap between current nutritional outcomes and both national and global targets. Nepal’s Second Long-Term Health Plan (1997–2017) and the Multi-Sector Nutrition Plans I and II aimed to reduce wasting to below 5%, a goal that remains unmet. Furthermore, the Sustainable Development Goals call for reducing wasting to less than 5% by 2025 and to 4% by 2030 [4].

To address acute malnutrition, the Government of Nepal introduced the Community-Based Management of Acute Malnutrition (CMAM) program in 2008/09, later scaled up as the Integrated Management of Acute Malnutrition (IMAM). Initially implemented in five districts, the program had expanded to 38 districts by 2020. In addition, a Comprehensive Nutrition-Specific Intervention Package has been implemented across all 77 districts [5]. Despite these initiatives, a significant proportion of malnourished children remain undetected and untreated, highlighting critical gaps in program reach and accessibility.

In Bajura district of Sudurpaschim Province, IMAM services are provided through 43 Outpatient Therapeutic Care Centres (OTCCs), one Inpatient Therapeutic Centre (ITC) at the district hospital, and one Nutrition Rehabilitation Centre (NRC) within the same facility. While service provision continues, no comprehensive evaluation of IMAM program coverage and access has been undertaken in the district. Existing data systems such as DHIS-2 capture only routine quantitative indicators such as admissions and discharges, but fail to provide insights into programmatic barriers, enablers, or implementation quality.

Understanding program coverage is essential, as the overall impact of nutrition interventions depends on both the proportion of affected children reached and the effectiveness of treatment provided [6]. The Semi-Quantitative Evaluation of Access and Coverage (SQUEAC) methodology provides a pragmatic approach to assessing these dimensions. This study applies the SQUEAC approach in Bajura to estimate the current coverage of the IMAM program, identify key barriers and facilitators of access, and propose actionable strategies to enhance service uptake.

## Method

This study employed a Semi-Quantitative Evaluation of Access and Coverage (SQUEAC) approach to evaluate the coverage and barriers to the Integrated Management of Acute Malnutrition (IMAM) program in Bajura district, Nepal. Bajura is a mountainous district in Sudurpaschim Province, where IMAM services are provided through 43 Outpatient Therapeutic Care Centres (OTCCs), one Inpatient Therapeutic Care Centre (ITCC), and one Nutrition Rehabilitation Centre (NRC). Despite the availability of these services, there has been limited documentation on their reach and accessibility.

SQUEAC is a mixed-method, three-stage methodology designed to identify program performance bottlenecks and estimate coverage.

The SQUEAC methodology comprises the following stages:

- **Stage I:** Identification of potential barriers and enablers using both routine data and collection of qualitative data
- **Stage II:** Hypothesis testing through targeted case-finding
- **Stage III:** Wide-area survey to produce a statistically valid coverage estimate

### Stage I

In this initial stage, the study team reviewed routine program data including IMAM admissions, discharge outcomes, and MUAC (Mid-Upper Arm Circumference) measurements from the DHIS-2 and IMAM registers 2.6.

Simultaneously, qualitative data were collected to explore underlying factors influencing access and uptake of IMAM services. Data collection included:

- Key Informant Interviews (KIIs)
- Focus Group Discussions (FGDs)
- Direct Observations

Respondents were purposively selected to ensure representation from various stakeholder groups, including caretakers of both covered and uncovered SAM children by the IMAM program, health workers, community leaders, school teachers, and Nutrition Focal Persons.

Qualitative data were analyzed using a Barriers, Boosters, and Recommendations (BBR) framework. Barriers and enabling factors were categorized into eight domains, six of which aligned with WHO’s health system building blocks, and two cross-cutting themes: community engagement and gender equality and social inclusion (GESI). Triangulation was achieved across data sources, collection methods, and geographic locations to ensure validity.

To strengthen the analysis, each identified barrier and booster was scored using:

- Simple scoring (equal weight for all items)
- Weighted scoring (importance-based scale of 1 to 4)

### Stage II

Based on insights from Stage I, a hypothesis was formulated:

*“Coverage of the program is high in areas/locations near the OTCC and low in areas/locations far”*

This hypothesis was tested through small-area surveys employing active and adaptive case-finding in rural areas and door-to-door screening in urban areas. Children aged 6–59 months were screened using MUAC and assessed for bilateral pitting oedema.

Ten locations were selected, i.e. four from rural areas (2 near and 2 far from OTCCs) and six from urban areas (3 near and 3 far from OTCCs). Standardized questionnaires adapted from the Coverage Monitoring Network (CMN) were used to collect data from:

Covered SAM cases: cases identified as SAM and enrolled in the treatment program during data collection

Uncovered SAM cases: identified as SAM during data collection but not enrolled in the treatment program

### Stage III

The third stage involved a Bayesian coverage estimation based on prior information from Stage I. The prior mode was calculated by averaging the simple and weighted scores from the BBR framework.

Coverage estimation used the following formula:

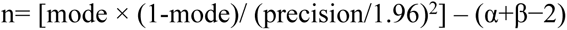

Where:

*mode: estimated prior coverage*

*precision: set at 15%*

*alpha and beta: set at 11.1 and 20.6 respectively (25% uncertainty level)*

*sample size (n): number of SAM cases to be identified*

The number of locations required was calculated using:

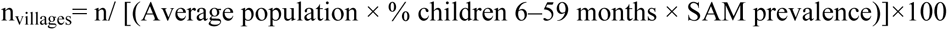

Parameters used:

*Average population per area: 604 (National Census, 2021)*

*Children under 5: 9.1% (HMIS 2023/24)*

*SAM prevalence by MUAC: 1.1% (SMART Nutrition Survey, Bajura 2022)*

Using this, 16 locations were sampled. Case identification was done using MUAC tapes and oedema check. Responses were logged using adapted CMN questionnaires and coverage tally sheets. The data collection period for the entire three stages ranged from 24 November to 06 December 2024.

## Ethical Statement

Ethical approval for this study was obtained from the Nepal Health Research Council (NHRC) under the approval number 543_2024. Participation in the study was voluntary, and informed written consent was obtained from all participants prior to interviews and focus group discussions. All data were anonymized to ensure participant privacy.

## Results

The results are presented by stages as this assessment is 3 phase procedures where the results of one stage lead to another.

**Stage I:** The routine data from HMIS for the year 2023/24 (2080/81 BS) was analyzed for admission over time, discharge over time and MUAC on admission, length of stay was analyzed form HMIS 2.6 (IMAM register) maintained at OTC centers.

### Quantitative Findings

#### Admission over time

Analysis of HMIS-reported data for fiscal year 2023/24 AD (2080/81 BS), retrieved from the DHIS2 platform, indicates that 179 children were admitted for treatment during the reporting period. Admissions were recorded year-round, with a notable rise between mid-April and mid-July 2024. This period coincides with the summer months and the onset of the monsoon season (mid-May to mid-June), when waterborne and vector-borne diseases such as diarrhoea and typhoid are more prevalent—potentially contributing to the observed increase.

The three-month moving median (M3A3) analysis revealed relatively stable admission numbers during the early months (mid-July to mid-October 2023), followed by a decline between mid-October 2023 and mid-January 2024. From mid-January onwards, admissions began to rise steadily, surpassing or matching the expected trend, and reaching their peak in mid-May to mid-June 2024.

**Figure 1:**
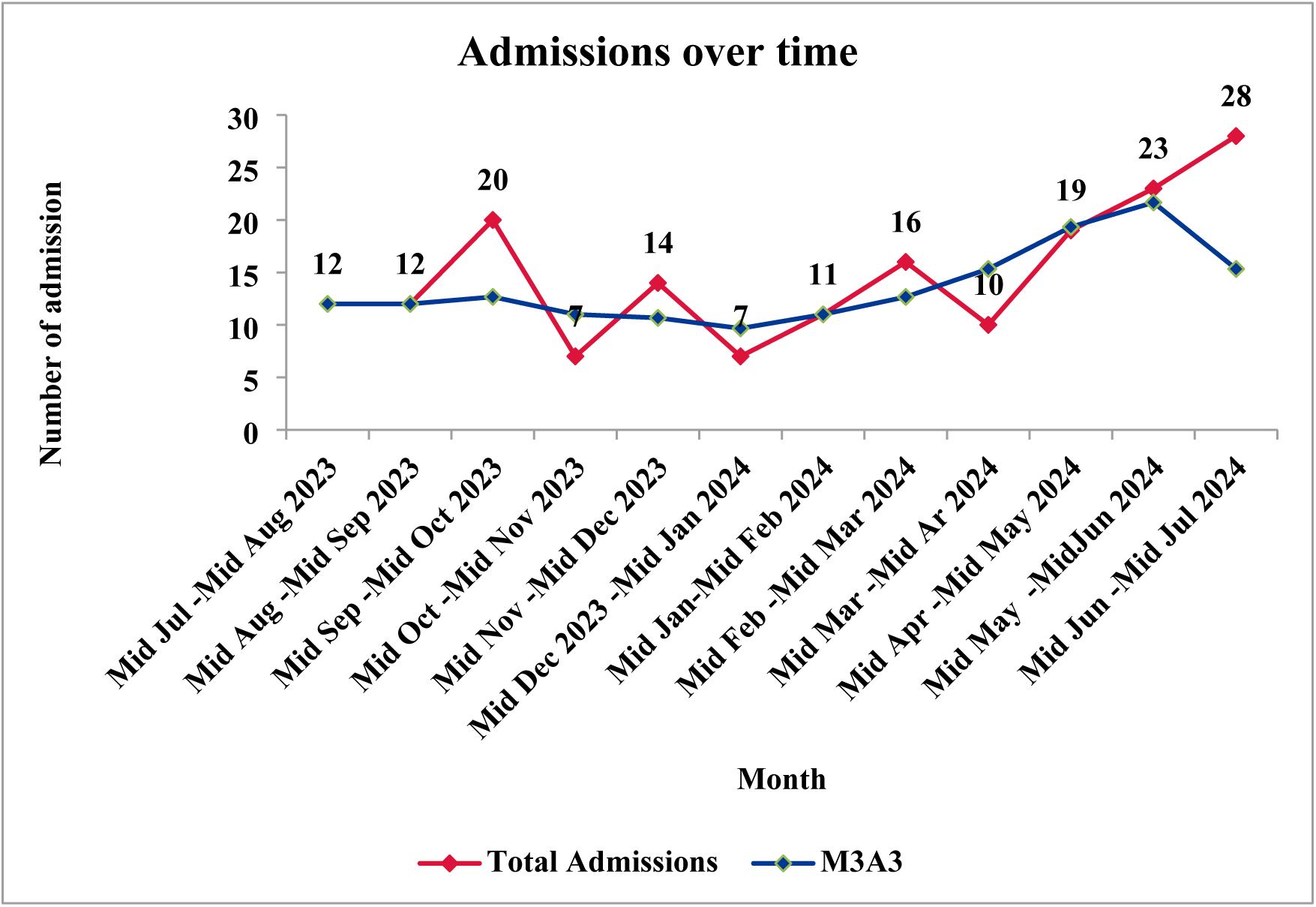
Admission over time.

### MUAC at admission

The data analysis on MUAC at admission reveals that 50% of the cases were admitted with a MUAC of 114 mm. However, there were instances where the MUAC values were not recorded in the register. The data also indicates that children with MUAC measurements greater than 115 mm, ranging from 115 to 125 mm, were admitted to the OTCC. Additionally, cases with MUAC measurements as low as 100 mm, 105 mm, and 110 mm were recorded. This highlights late detection and delayed admission of cases into the program. Such delays place children at a significantly higher risk of health complications and vulnerability. The admission at MUAC > or equal to 115 is because in the context of Nepal, admission in OTCC is done by z-score or MUAC criteria. In few cases, it might be because of wrong admission.

**Figure 2:**
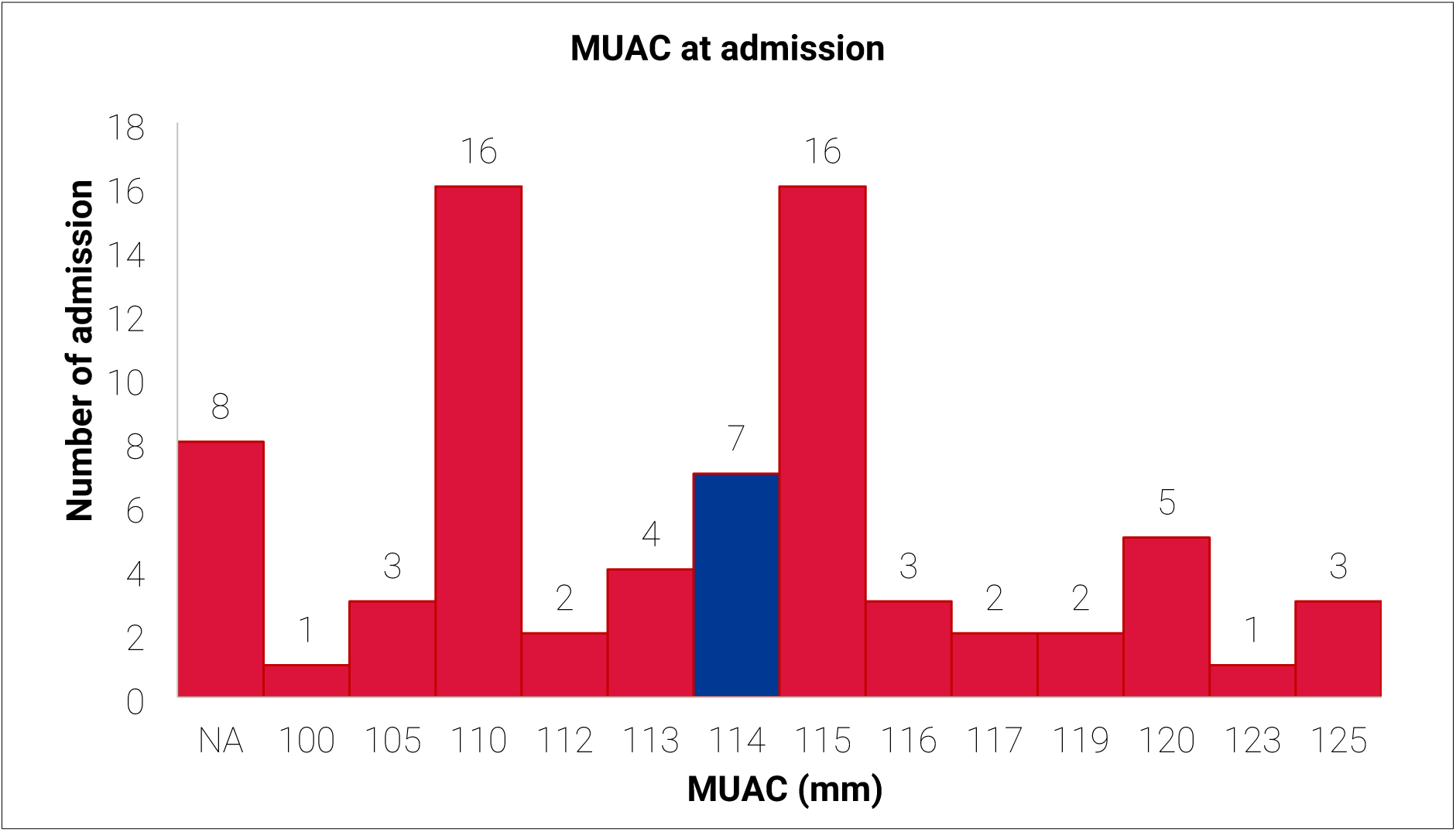
MUAC at admission.

### Discharge over time

Discharge outcomes for children include those discharged as recovered, defaulted or dead. A well- performing program should achieve a recovery rate of >75%, a death rate of <10%, and a default rate of <15%, according to the Sphere standards.

**Figure 3:**
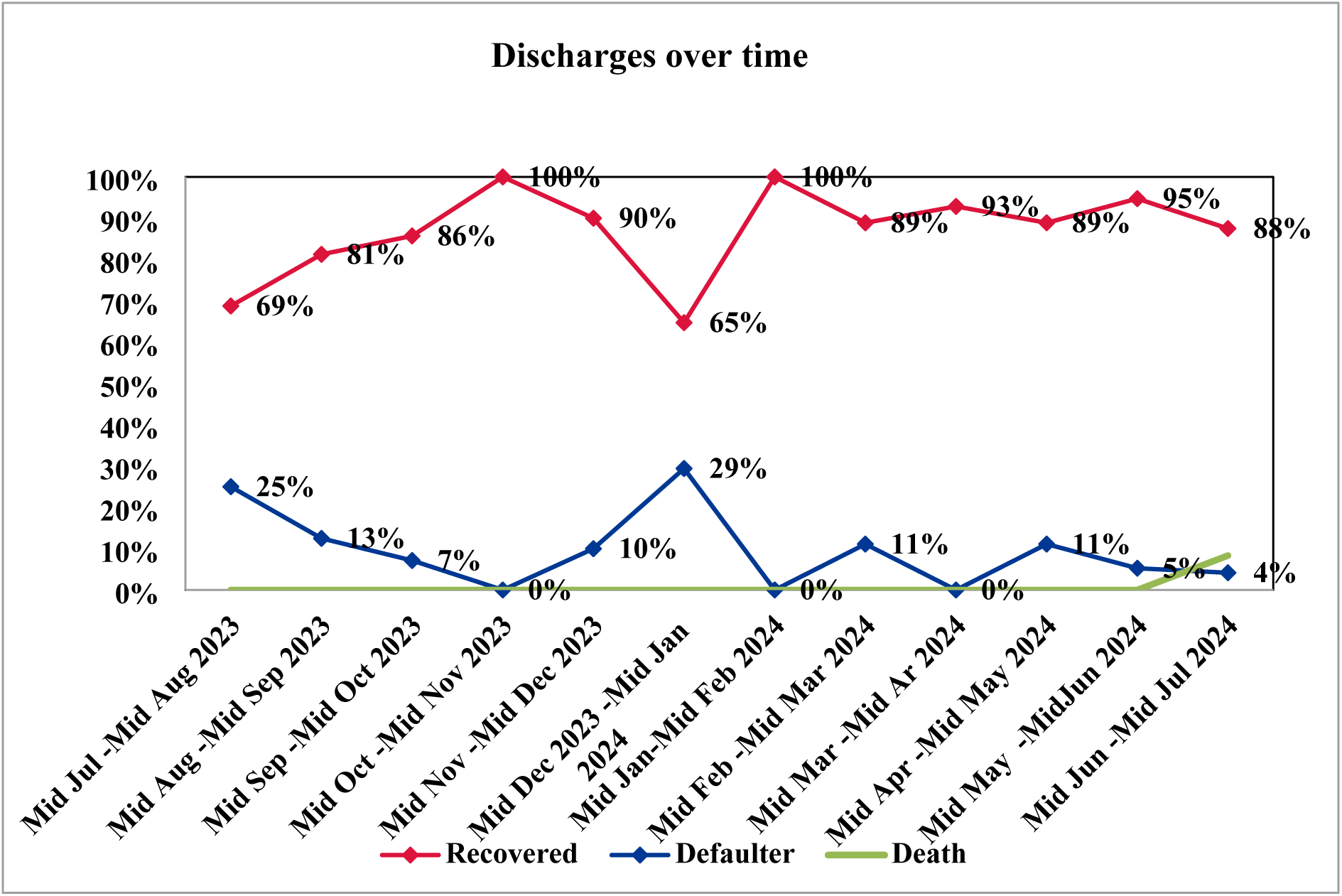
Discharges over time.

However, in the months of Mid-July to Mid-August, the recovery rate falls below the Sphere standard. This may be attributed to the monsoon season, during which the prevalence of diseases is high. Additionally, monsoon-related challenges, such as landslides and rising riverbeds, can hinder access to health services in the hills and mountains, potentially contributing to the lower recovery rates. Similarly, in the month of Mid-December 2023 to Mid-January 2024, the recovery rate is below the Sphere standard, possibly due to peak winter conditions, including snowfall, which restrict mobility and access to health facilities. The defaulter rate during these months is notably high, exceeding the Sphere standard. For instance, the defaulter rate in Mid-July to Mid-August is 25%, while in Mid-December to Mid-January, it rises to 29%. Cases of children being discharged as not improved were also observed in the district, further highlighting gaps in achieving optimal outcomes.

### Length of stay

**Figure 4:**
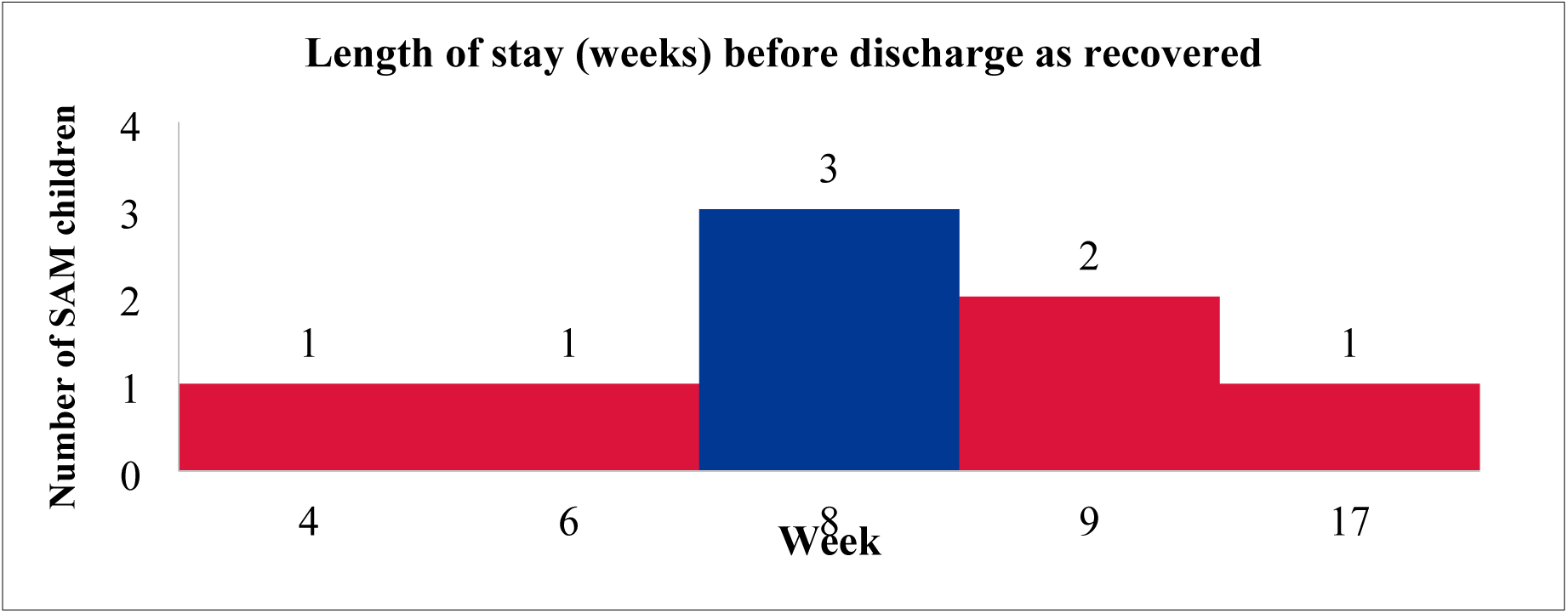
Length of stay.

Data for length of stay of the recovered children was analysed by compiling the data of HMIS register 2.6 maintained at different OTCCs. The figure below shows that 50% of children enrolled or admitted as SAM were recovered within 8 weeks of treatment. The length of stay for recovery ranges from a minimum of 4 weeks to a maximum of 17 weeks of treatment.

### Length of stay before defaulting

The graph below, based on data from HMIS 2.6 from 10 different OTCCs in Bajura district, highlights defaulting trends among children enrolled in the program. It reveals that 50% of children defaulted after attending only one visit, meaning they did not return for any follow-up. Additionally, the data indicates that the remaining 50% defaulted at various stages after their first visit, with the number of visits before defaulting ranging from 2 to 6.

**Figure 5:**
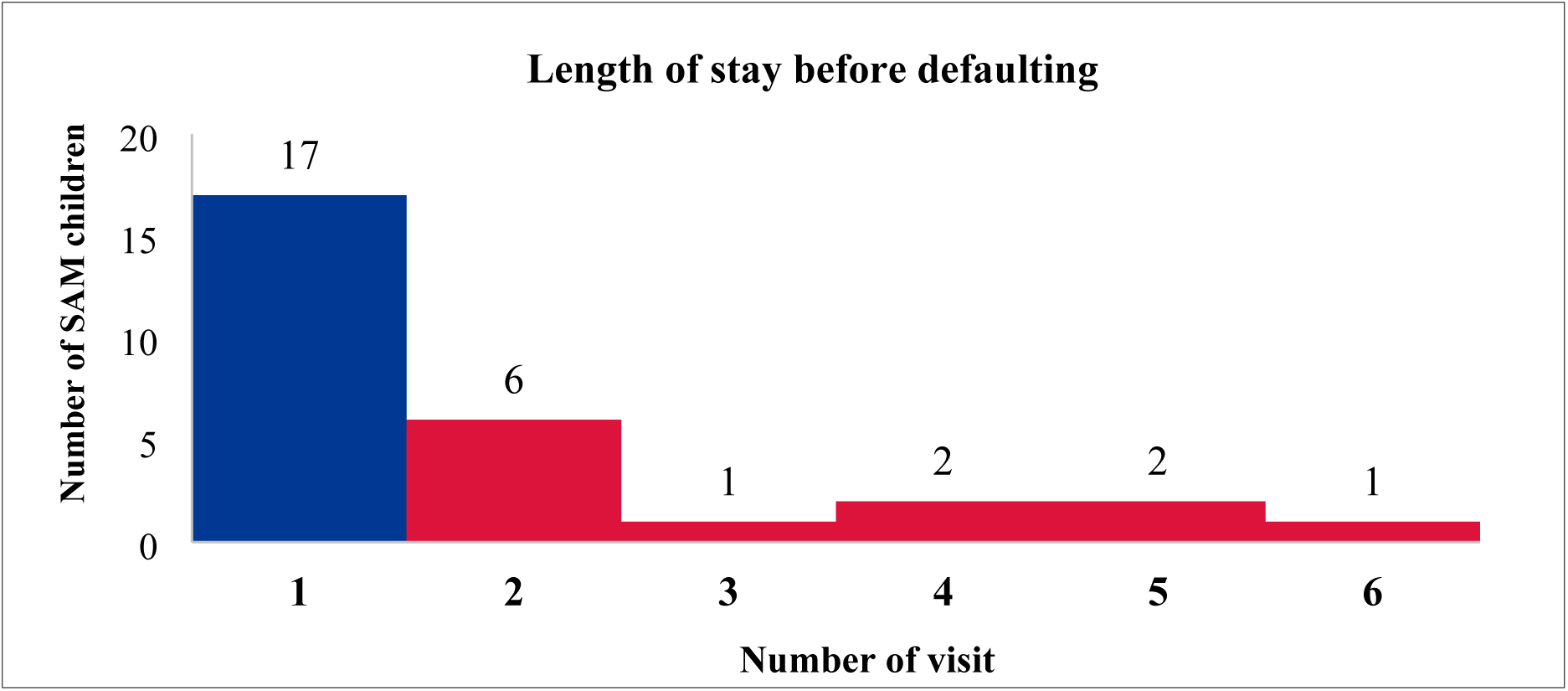
Length of stay before defaulting.

The qualitative data collection from 14 locations identified 14 boosters and 25 barriers affecting the coverage and access of the program. Boosters primarily involved service-related factors such as the expansion of OTCCs, availability of IMAM guidelines (not updated/recent), integration of screening into Growth Monitoring and Promotion (GMP), and immunization platforms, effective follow-up systems, regular monthly screenings by FCHVs, positive health workers’ behavior, strong coordination between FCHVs and OTCCs/health facilities, and coordination with ward offices for health promotion activities like Mothers’ Group Meetings (MGMs), food demonstrations, and *"Poshan Saptaha"* (Nutrition Week) events in selected schools.

Despite having boosters that helped for reaching SAM children and enrolling them into IMAM program, it faced several barriers particularly in the unavailability of updated guidelines and IEC materials in functional OTCCs, inadequate recording tools (IMAM cards, registers, and referral slips), incomplete monthly recording and reporting in the IMAM registers at OTCCs, limited/lack of training and refreshers to health workers, insufficient human resources, inactive or non-functional newly established OTCCs, and frequent stockouts of RUTF at OTCCs.

### Concept Mapping

Each barrier and booster were systematically analysed to understand its underlying causes and interconnections. The analysis revealed that barriers were not isolated; rather, they were interlinked, with one barrier reinforcing or leading to another. A similar pattern was observed among boosters, where positive factors contributed to and reinforced each other. The concept mapping below illustrates the relationship between the identified barriers and boosters.

**Figure 6:**
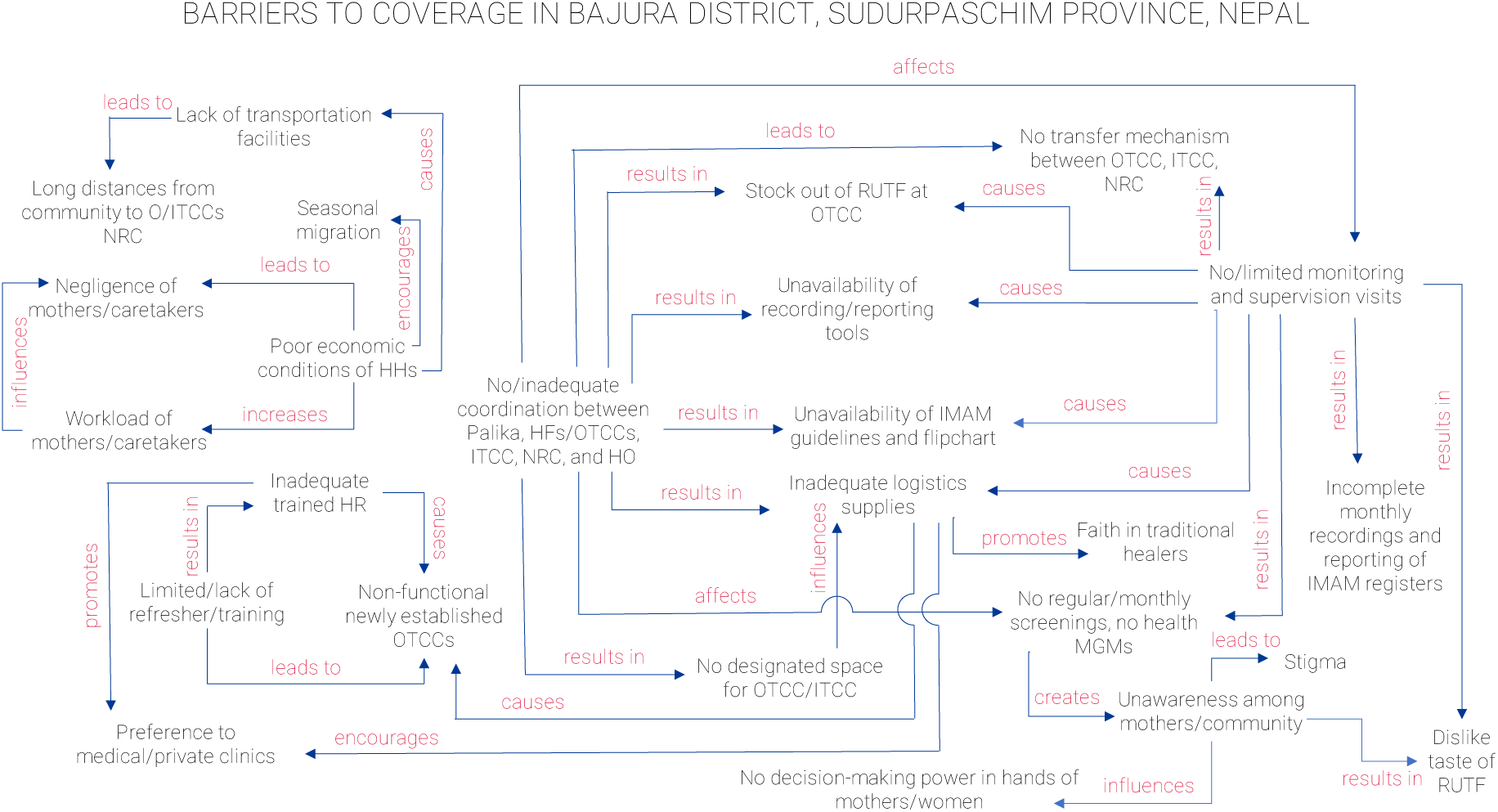
Barriers to coverage in Bajura.

**Figure 7:**
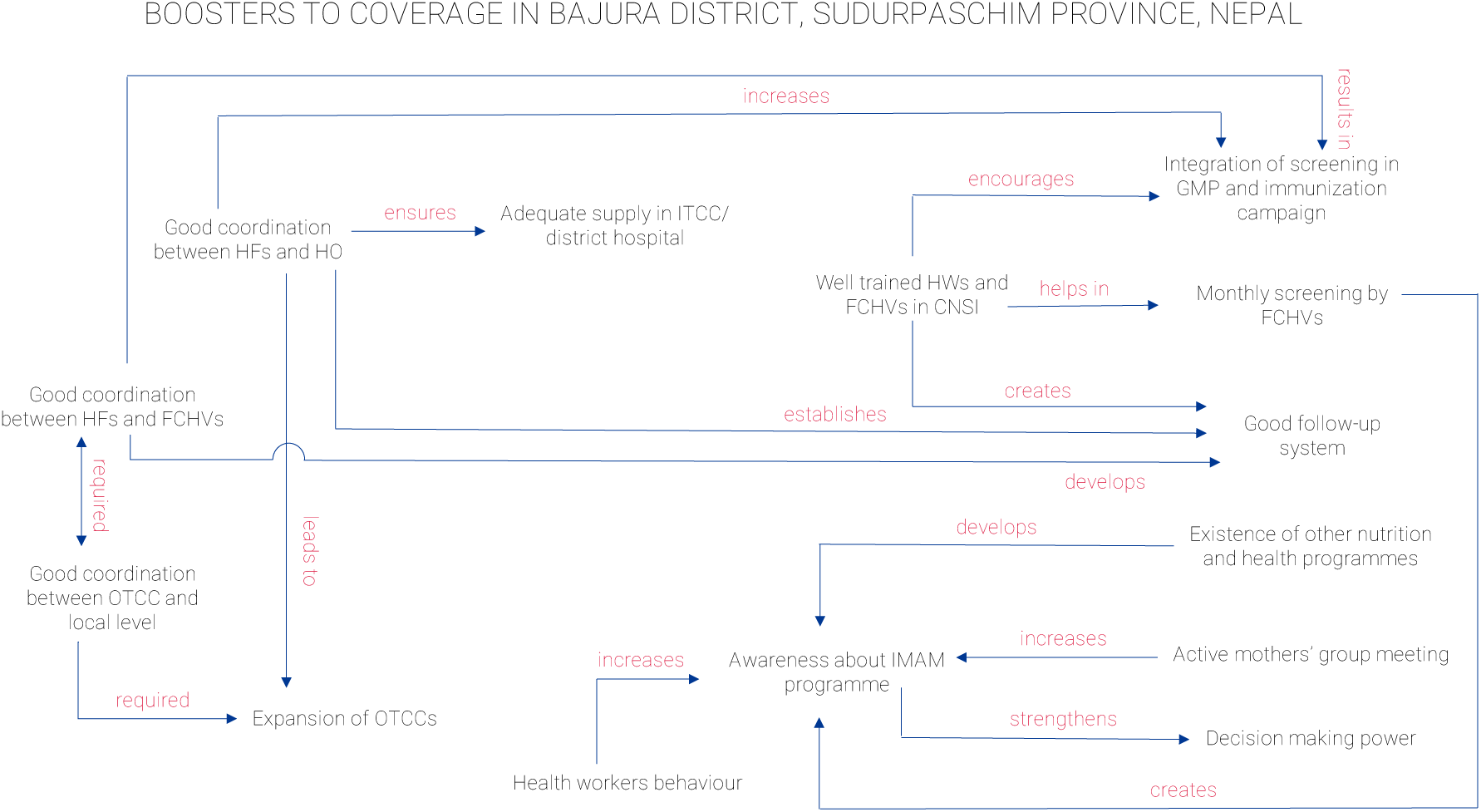
Boosters affecting Coverage of the program.

The BBQ exercise evaluated 14 boosters and 25 barriers using both a simple scoring and a weighted scoring method. In the simple scoring method, each booster and barrier were assigned an equal score, assuming all factors have equal influence on the program’s effectiveness. In contrast, the weighted scoring method assigned scores from 1 to 4 based on the perceived level of impact, each factor has on the program’s outcomes.

### Stage II

Based on the quantitative data and qualitative data collection and analysis, assessment team developed and tested following hypothesis in both rural and urban context.

Hypothesis: ***“Coverage of the program is high in areas/locations near the OTCC and low in areas/locations far”***

Here, the distance was defined as near if it takes less or equal to 30 minutes by walking or by any other means of transportation and far if it takes more than 30 minutes by walking or by any other transportation means to reach the OTCC to seek service delivery.

In the rural context, decision rule for hypothesis testing validation is decision (d)=n/2 and for the urban context, d=70% of n. Here, n is number of total numbers of SAM cases identified during the survey. If the number of covered cases in near locations is greater than decision rule, the hypothesis is confirmed and if the number of covered cases in farther locations is less than decision rule, the hypothesis is confirmed.

**Table 1:** Hypothesis testing and validation in rural context.

| Rural context |  |  |  |
| --- | --- | --- | --- |
| High coverage: | Calculation of decision rule / result |  | Validation of hypothesis |
| Total SAM cases: 3 | Target coverage = 50% |  |  |
| Covered cases: 3 | $n = 3$ | Number of covered cases<br>(3) > decision rule (1) | ***Hypothesis is confirmed*** |
| | $d = n/2$ | | |
|  | 1.5 |  |  |
|  | 1 |  |  |
| Low coverage: | Target coverage = 50% |  |  |
| Total SAM cases: 2 | $n = 2$ | number of covered cases<br>(0) < decision rule (1) | ***Hypothesis is confirmed*** |
| Covered case:0 | $d = n/2$ | | |
|  | 1 |  |  |

**Table 2:**
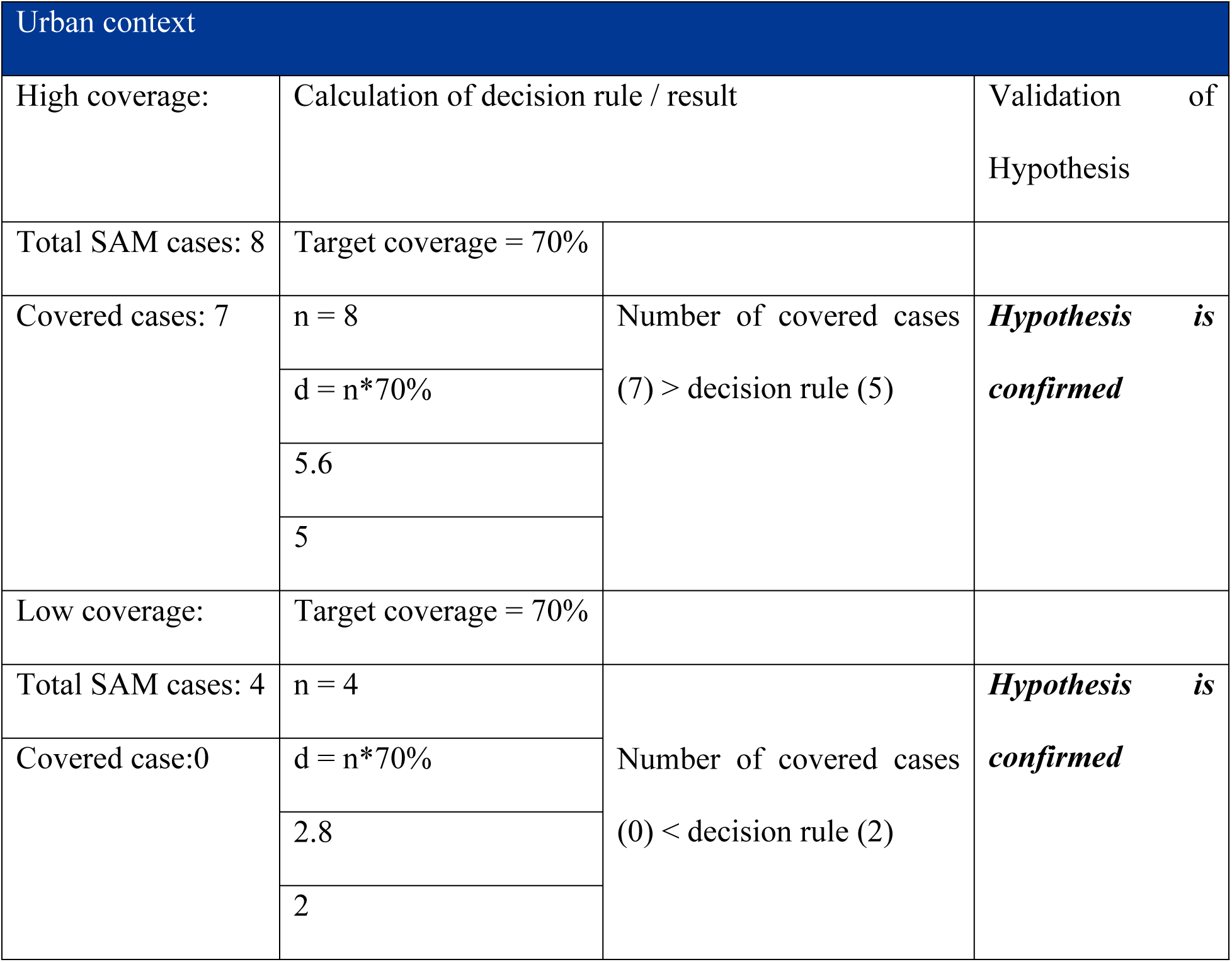
Hypothesis testing and validation in urban context.

| Urban context |  |  |  |
| --- | --- | --- | --- |
| High coverage: | Calculation of decision rule / result |  | Validation of Hypothesis |
| Total SAM cases: 8 | Target coverage = 70% |  |  |
| Covered cases: 7 | n = 8 | Number of covered cases<br>(7) > decision rule (5) | ***Hypothesis is confirmed*** |
|  | d = n*70% |  |  |
|  | 5.6 |  |  |
|  | 5 |  |  |
| Low coverage: | Target coverage = 70% |  |  |
| Total SAM cases: 4 | n = 4 | Number of covered cases<br>(0) < decision rule (2) | ***Hypothesis is confirmed*** |
| Covered case:0 | d = n*70% |  |  |
|  | 2.8 |  |  |
|  | 2 |  |  |

In both rural and urban context hypothesis was confirmed and hence the stage 3 or wide area is proceeded.

### Stage III

The sample size of 10 SAM cases to be identified from 16 different locations. However, a total of 26 SAM cases were identified, of which only three were enrolled in the program, highlighting a significant gap in program coverage. Notably, the majority of the identified cases (18 out of 26) were female children, suggesting a potential gender-related disparity in nutritional status or access to care.

Among 23 uncovered cases, 15 were female children, and 8 were male children. This indicates that female children represented the majority of SAM cases. These findings point to the need for targeted interventions to address barriers to program access for female children, and to promote greater awareness and utilisation of available services.

Interviews with caretakers of uncovered cases revealed three primary barriers. First, stockouts of Ready-to-Use Therapeutic Food (RUTF) during follow-up visits discouraged return, highlighting the need for uninterrupted supply to sustain caretaker trust and service utilization. Second, among 23 uncovered severe acute malnutrition (SAM) cases, eight had previously enrolled but discontinued due to children’s dislike of RUTF taste and stockouts, underscoring challenges in both acceptance and treatment continuity. Third, lack of awareness about the program and malnutrition led some caretakers to forgo enrolment, often perceiving their children as healthy. These findings point to the importance of ensuring consistent supply, improving RUTF acceptability, and strengthening community engagement to enhance coverage and retention. With the number of cases found during wide area survey coverage was estimated. Among 26 cases, covered cases (C_in_ )=3 and Uncovered cases (C_out_) = 23. Using Bayes calculator, the conjugate analysis was done with the prior parameters for Prior estimate of 35.1%. The overall coverage estimate is found to be 23.5% (14.5% - 36.0%) with z=1.97 and p-value =0.0486. The p-value (0.0486) shows that the observed data differs slightly from the prior assumptions, but they are not completely unrelated. This means the new data has updated our understanding and influenced the posterior distribution providing a more accurate picture of the program coverage by combining the prior knowledge with the new evidence generated in the likelihood or wide area survey.

**Figure 8:**
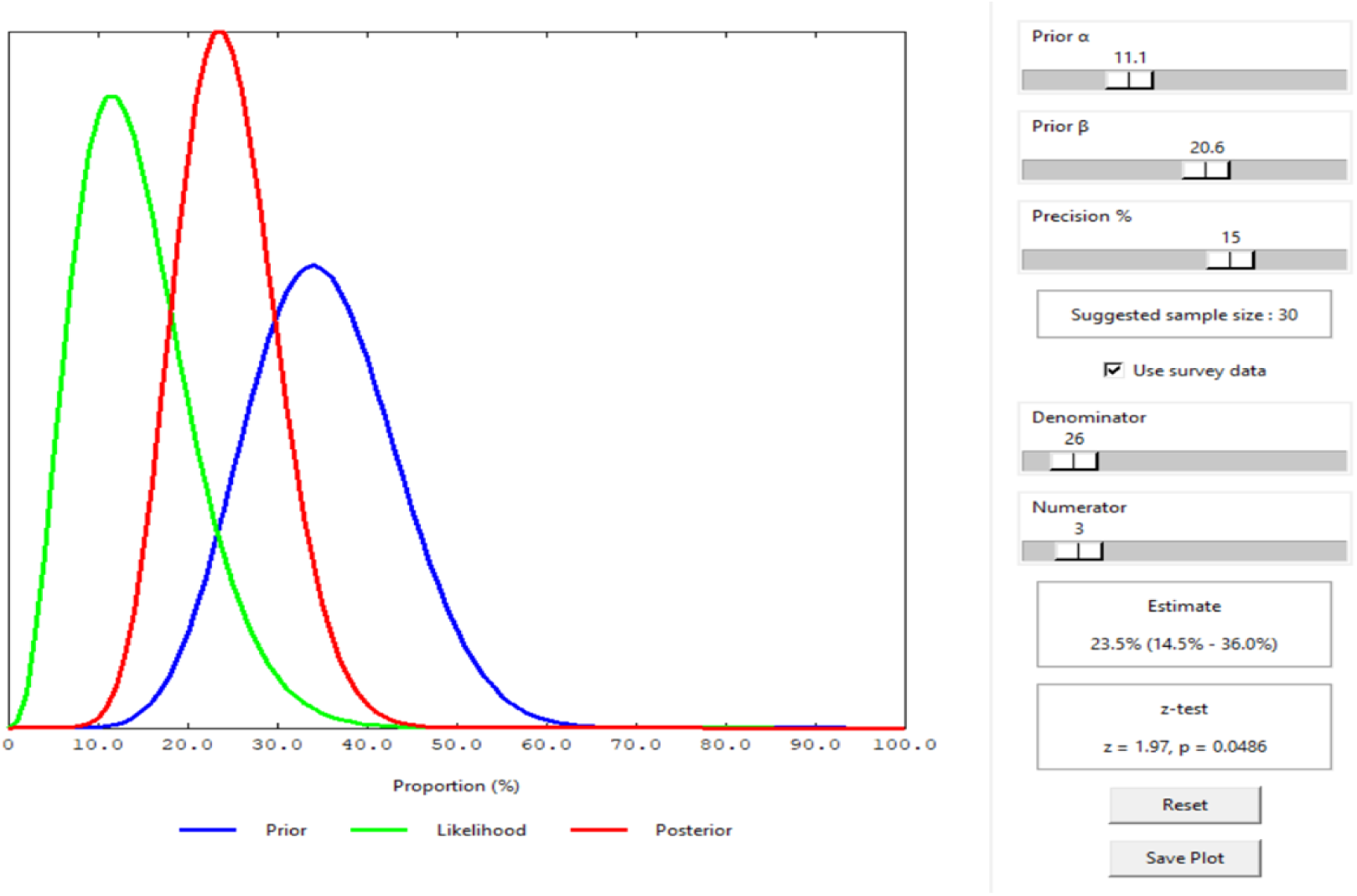
Coverage estimation using Bayesian scale.

The Bayes calculator provides the overall coverage estimate. The point coverage of the district is calculated using following formula.

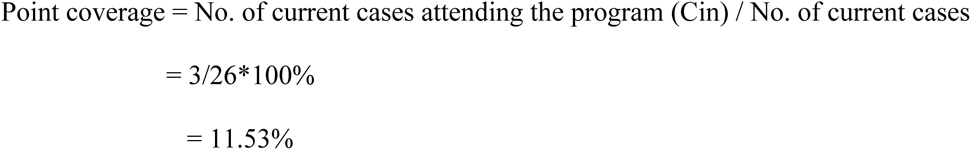

Since, no recovering cases was found in the case of Bajura, period coverage could not be calculated.

## Discussion

The IMAM program for children under five years of age has been operational in Bajura district since 2014/15. Despite nearly a decade of implementation, there has been no systematic evidence generated on program access and coverage, apart from routine program data capturing enrolment of cases and discharge outcomes. Such routine data, while valuable, does not provide a comprehensive picture of how well the program is reaching its target population or the barriers faced by caretakers. For impactful and equitable implementation, periodic coverage investigations together with an understanding of the factors influencing coverage are essential to improving both reach and effectiveness, thereby ensuring the program meets identified needs (7). This coverage assessment was undertaken to address this evidence gap, with the dual objective of identifying areas of high and low coverage and providing an overall estimate of program coverage.

The overall coverage estimate of 23.5% (95% CI: 14.5–36.0%), derived using Bayesian analysis, highlights considerable gaps compared to the prior estimate of 35.1%. These gaps appear to stem from multiple socio-cultural barriers, such as geographical inaccessibility, misconceptions among mothers and caretakers, and limited transportation options (8). On the service delivery side, challenges included a preference among some caregivers for medical or private clinics, unavailability of essential supplies in health facilities particularly stockouts of RUTF, an inadequate number of trained health workers, poor record-keeping systems, and the absence of dedicated spaces for treatment at OTCCs (9). Each of these factors directly or indirectly undermines the program’s ability to identify, enroll, and retain children in treatment.

Despite these challenges, there have been notable programmatic improvements, including the expansion of OTCCs and the integration of SAM screening into routine growth monitoring and immunization sessions, which have supported earlier detection of cases. Nevertheless, the point coverage reflecting the proportion of currently enrolled cases was found to be just 11.53%. This very low figure signals the program’s limited reach at any given time and underscores the urgency of implementing innovative, context-specific strategies to strengthen community-level case identification, increase enrolment, and ensure continuity of care for children affected by severe acute malnutrition.

## Conclusion

Strengthening the existing health system is critical to improving coverage, access, and meeting needs. Key priorities include building the capacity of health workers, ensuring a consistent supply of RUTF, and investing in community engagement to raise awareness about malnutrition. Equally important are ensuring the full functionality of newly established OTCCs, providing regular supportive supervision, and establishing strong feedback mechanisms. Improving data management systems, adopting data-driven approaches, and enhancing service accessibility, particularly for remote communities will be essential to achieving sustained program impact.

## Data Availability

All relevant data are within the manuscript and its supporting information files

